# Viral RNA level, serum antibody responses, and transmission risk in discharged COVID-19 patients with recurrent positive SARS-CoV-2 RNA test results: a population-based observational cohort study

**DOI:** 10.1101/2020.07.21.20125138

**Authors:** Chao Yang, Min Jiang, Xiaohui Wang, Xiujuan Tang, Shisong Fang, Hao Li, Le Zuo, Yixiang Jiang, Yifan Zhong, Qiongcheng Chen, Chenli Zheng, Lei Wang, Shuang Wu, Weihua Wu, Hui Liu, Jing Yuan, Xuejiao Liao, Zhen Zhang, Yiman Lin, Yijie Geng, Huan Zhang, Huanying Zheng, Min Wan, Linying Lu, Xiaohu Ren, Yujun Cui, Xuan Zou, Tiejian Feng, Junjie Xia, Ruifu Yang, Yingxia Liu, Shujiang Mei, Baisheng Li, Zhengrong Yang, Qinghua Hu

## Abstract

**Summary:** *Background:* Managing discharged COVID-19 (DC) patients with recurrent positive (RP) SARS-CoV-2 RNA test results is challenging. We aimed to comprehensively characterize the viral RNA level and serum antibody responses in RP-DC patients and evaluate their viral transmission risk.

*Methods:* A population-based observational cohort study was performed on 479 DC patients discharged from February 1 to May 5, 2020 in Shenzhen, China. We conducted RT-qPCR, antibody assays, neutralisation assays, virus isolation, whole genome sequencing (WGS), and epidemiological investigation of close contacts.

*Findings:* Of 479 DC patients, the 93 (19%) RP individuals, including 36 with multiple RP results, were characterised by young age (median age: 34 years, 95% confidence interval [CI]: 29–38 years). The median discharge-to-RP length was 8 days (95% CI: 7–14 days; maximum: 90 days). After readmission, RP-DC patients exhibited mild (28%) or absent (72%) symptoms, with no disease progression. The viral RNA level in RP-DC patients ranged from 1·9–5·7 log_10_ copies/mL (median: 3·2, 95% CI: 3·1–3·5). At RP detection, the IgM, IgG, IgA, total antibody, and neutralising antibody (NAb) seropositivity rates in RP-DC patients were 38% (18/48), 98% (47/48), 63% (30/48), 100% (48/48), and 91% (39/43), respectively. Regarding antibody levels, there was no significant difference between RP-DC and non-RP-DC patients. The antibody level remained constant in RP-DC patients pre- and post-RP detection. Virus isolation of nine representative specimens returned negative results. WGS of six specimens yielded only genomic fragments. No clinical symptoms were exhibited by 96 close contacts of 23 RP-DC patients; their viral RNA (96/96) and antibody (20/20) test results were negative. After full recovery, 60% of patients (n=162, 78 no longer RP RP-DC and 84 non-RP-DC) had NAb titres of ≥1:32.

*Interpretation:* RP may occur in DC patients following intermittent and non-stable excretion of low viral RNA levels. RP-DC patients pose a low risk of transmitting SARS-CoV-2. An NAb titre of ≥ 1:32 may provide a reference indicator for evaluating humoral responses in COVID-19 vaccine clinical trials.

*Funding:* Sanming Project of Medicine in Shenzhen, China National Science and Technology Major Projects Foundation, Special Foundation of Science and Technology Innovation Strategy of Guangdong Province of China, and Shenzhen Committee of Scientific and Technical Innovation grants.

## Introduction

Coronavirus disease 2019 (COVID-19), caused by severe acute respiratory syndrome coronavirus 2 (SARS-CoV-2), has spread globally to over 213 countries.^1-5^ As of July 10, 2020, there have been more than 12,000,000 confirmed patients and 540,000 deaths. Currently, there are approximately 200,000 new confirmed patients daily, posing huge challenges for public health and medical institutions.

Worldwide, there are more than 6,500,000 recovered COVID-19 patients.^4^ Recent reports have described discharged COVID-19 (DC) patients with recurrent positive (RP) reverse transcription quantitative PCR (RT-qPCR) test results for SARS-CoV-2 (RP-DC patients).^6-10^ These studies focused on the clinical characteristics of a small number (<40) of RP-DC patients and found that they generally showed no clinical symptoms or disease progression. However, their positive SARS-CoV-2 RNA test results suggest that these patients might be virus carriers. The management of RP-DC patients is challenging because of the current lack of understanding regarding their viral RNA level, antibody responses, and viral transmission risk. In China, RP-DC patients are placed under a costly fourteen-day quarantine. Clarifying the characteristics and viral transmission risk of RP-DC patients is critical for appropriately managing their cases.

We performed a population-based observational cohort study of 479 DC patients, discharged from February 1 to May 5, 2020 in Shenzhen, China. Based on the results of integrating RT-qPCR, antibody assays, neutralisation assays, virus isolation, whole genome sequencing (WGS), and epidemiological investigation of close contacts, we comprehensively detailed the demographic, clinical, viral RNA level, and antibody response characteristics and evaluated the viral transmission risk of RP-DC patients.

## Methods

### Patients

All COVID-19 patients in Shenzhen were treated at the designated Shenzhen Third People’s Hospital; their cases were reported to Shenzhen Center for Disease Control and Prevention (CDC)^11^. This study enrolled all DC patients discharged from February 1 to May 5, 2020 in Shenzhen, including asymptomatic patients identified during the RT-qPCR screening of confirmed COVID-19 patient close contacts (Figure 1a). Discharge criteria included: (1) normal temperature for >3 days, (2) resolved respiratory symptoms, (3) substantial pulmonary lesion absorption on chest computed tomography (CT) images, and (4) negative results from two consecutive SARS-CoV-2 RNA tests conducted >1 day apart. After discharge, DC patients were quarantined at home (before February 18) or in centralised facilities (from February 18) for 14 days. During the 14-day quarantine period, both nasopharyngeal and anal swabs (n=2,442, 4–20 per person) were collected from each patient on the 7^th^ and 14^th^ days (before March 18) or the 1^st^, 3^rd^, 7^th^, and 14^th^ days (from March 18) for SARS-CoV-2 RNA detection by RT-qPCR. From March 18, serum specimens were collected on the 1^st^, 3^rd^, 7^th^, and 14^th^ days for antibody assays (n=499, 2–8 per person), and some RP-DC patient blood specimens (n=147, 1–4 per person) were collected for SARS-CoV-2 RNA detection by RT-qPCR. After quarantine, DC patients were regularly followed-up on the 7^th^, 14^th^, 30^th^, and 60^th^ days post-discharge. Demographic and clinical severity information was extracted from electronic hospital medical records. Clinical severity on first admission was classified as asymptomatic, mild, moderate, or critical based on Chinese Guidelines for Diagnosis and Treatment for Novel Coronavirus Pneumonia^12^.

**Figure 1.**
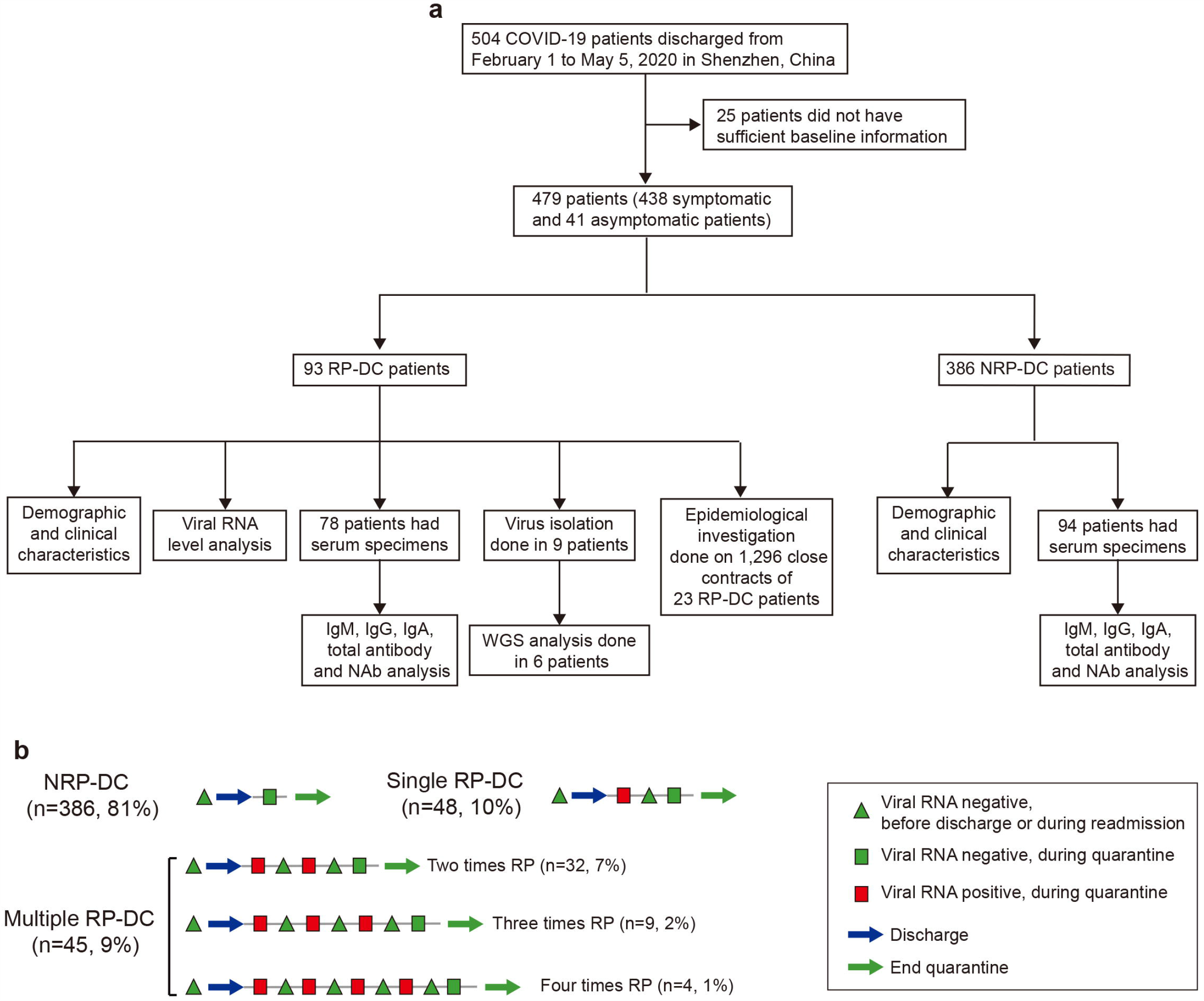
(a–b) Profile of the discharged COVID-19 patients included in this study (a) and case definition concept figure (b).

The study was approved by the Ethics Committee of Shenzhen CDC (QS2020060007). As data collection is part of the public health investigation of an emerging outbreak, individual informed consent was waived.

### Case definition

Because negative results from two consecutive SARS-CoV-2 RNA tests were part of the discharge criteria, a DC patient with recurrent positive test results was defined as an RP-DC patient (Figure 1b and appendix Figure S1). These patients were readmitted to hospital for further medical observation until they met the discharge criteria again, including negative results from two consecutive SARS-CoV-2 RNA tests. After re-discharge, an RP-DC patient with further positive SARS-CoV-2 RNA test results was defined as a multiple-RP-DC patient. A DC patient with constant negative SARS-CoV-2 RNA test results was defined as a non-RP-DC (NRP-DC) patient.

### Procedures

SARS-CoV-2 RT-qPCR tests were performed on the day of sampling using commercial kits (Zhongshan Daan Biotech). After 45 cycles, specimens with cycle threshold (Ct) values of ≤40 for both tested genes were considered positive; single-gene-positive specimens were retested and considered positive if the Ct values from the repeat tests were ≤ 40. The viral RNA level (copies/mL) was calculated from Ct values based on the standard curve of control product (Zhongshan Daan Bio-Tech, appendix Figure S2). Serum immunoglobulin (Ig) antibody against the SARS-CoV-2 surface spike protein receptor-binding domain (RBD) was measured using a chemiluminescence kit (IgM, IgG, and total antibody, Beijing Wantai Biotech, measured by cut-off index [COI]) or enzyme-linked immunosorbent assay kit (IgA, Beijing Hotgen Biotech, measured by optical density at 450/630 nm [OD_450/630_]) in accordance with the manufacturer’s instructions. Virus neutralisation assays were performed using SARS-CoV-2 virus strain 20SF014/vero-E6/3 (GISAID accession number EPI_ISL_403934) in biosafety level 3 (BSL-3) laboratories to obtain the neutralising antibody (NAb) titre. To define the cut-off for seropositivity, 169 and 128 serum specimens from confirmed COVID-19 patients and healthy persons were used as positive and negative controls, respectively. Specimens with COI>1 (IgM, IgG, or total antibody), OD_450/630_>0.3 (IgA), or an NAb titre of ≥1:4 were considered positive. Vero-E6 cells were used for virus isolation in a BSL-3 laboratory. WGS was performed after specifically amplifying SARS-CoV-2 RNA. Epidemiological investigations were conducted on 96 close contacts (unprotected exposure) of 23 RP-DC patients, identified during follow-up. Detailed methods are provided in the Supplementary Appendix.

### Statistical analysis

We performed statistical analyses using R version 3.6.1. Categorical and continuous variables were compared using Chi-squared and Mann-Whitney *U* tests, respectively. Correlations were assessed using Spearman’s correlation test. For all tests, *p*<0·05 was considered statistically significant.

## Results

From February 1 to May 5, 2020, 504 COVID-19 patients were discharged in Shenzhen. We excluded 25 of them from this study because of insufficient baseline information and enrolled the remaining 479 (438 symptomatic and 41 asymptomatic) patients (Figure 1a). As of July 10, 93 (19%) RP-DC patients were identified, including 45 (9%) multiple-RP-DC patients with two (n=32, 7%), three (n=9, 2%), or four (n=4, 1%) RP results post-discharge (Figure 1b and appendix Figure S1). Of the 93 RP-DC patients, 70 (75%) were identified during their fourteen-day quarantine, and the remaining 23 (25%) were identified during follow-up. The median time from discharge to the first RP was 8 days (95% confidence interval [CI]: 7–14 days; maximum: 90 days). The median times from discharge to final RP and from disease onset to final RP (viral RNA duration time) were 15 days (95% CI: 9–21 days; maximum: 90 days) and 46 days (95% CI: 38–53 days; maximum: 113 days), respectively (Table 1, Figure 2a–b and appendix Figure S1).

**Table 1.**
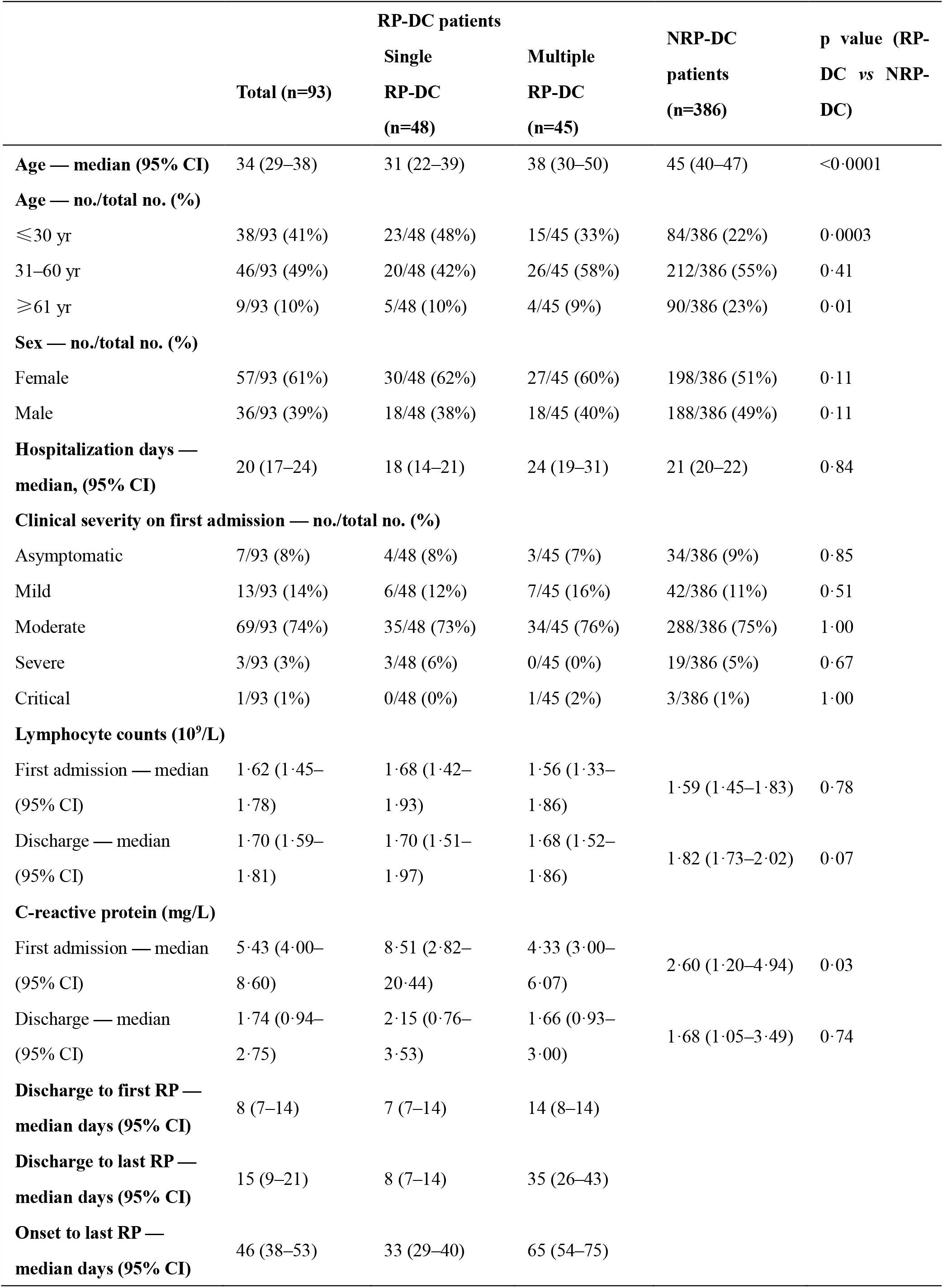
Demographic and clinical characteristics of RP-DC and NRP-DC patients.

**Figure 2.**
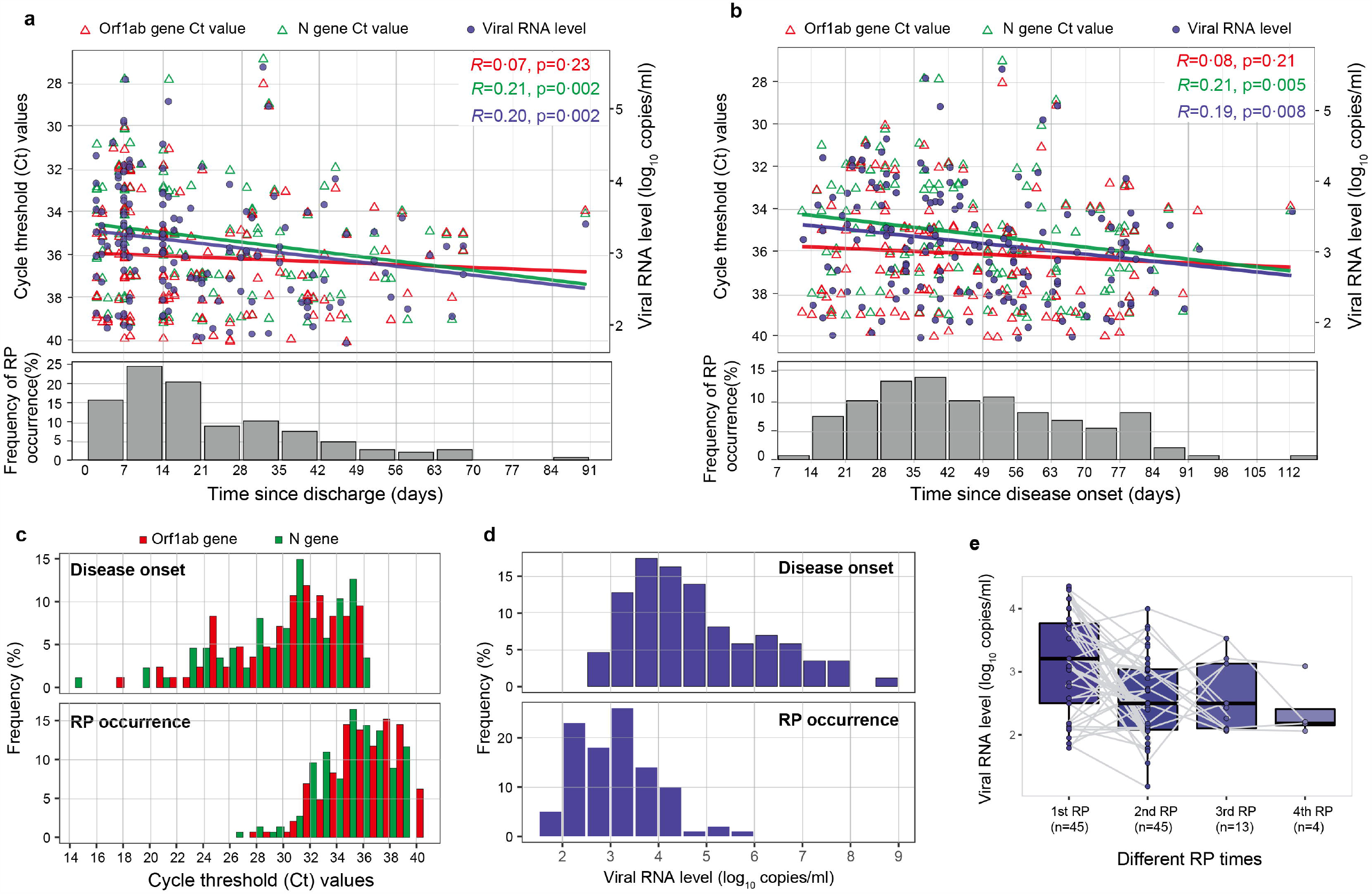
RT-qPCR cycle threshold (Ct) values and viral RNA levels in RP-DC patients. (a, b) Temporal distribution of Ct values (red and green triangles indicate the Orf1ab and N genes, respectively) and viral RNA levels (blue points) since discharge (a) or disease onset (b). The frequency of RP occurrence is shown by grey bars. (c) Ct values of RP-DC patients at the time of disease onset (top) or RP occurrence (bottom); colours indicate different target SARS-CoV-2 genes. (d) Estimated viral RNA level based on the correlation between viral RNA level and Ct value at the time of disease onset (top) or RP occurrence (bottom). (e) Viral RNA level dynamics in multiple-RP-DC patients. Specimens from individual patients are linked by grey lines.

There were more female (57/93, 61%) than male RP-DC patients (36/93, 39%, Table 1). This group was significantly younger (median age: 34 *vs* 45 years, *p*<0·0001) compared with the NRP-DC patients, with 41% of RP-DC patients aged under 30 years *vs* 22% of NRP-DC patients (*p*=0·0003). RP-DC patients had a median hospitalization period of 20 days, and their clinical severity on first admission was mostly moderate (69/93, 74%) or mild (13/93, 14%). No RP-DC patients had underlying immunodeficiency diseases, and 14 RP-DC patients (15%) were treated with steroids (methylprednisolone and/or dexamethasone) during hospitalization. There were no significant differences between RP-DC and NRP-DC patients in terms of hospitalization period, clinical severity on first admission, or steroid use (*p*>0·05). The C-reactive protein (CRP) level of RP-DC patients on first admission was significantly higher than that of NRP-DC patients (*p*=0·03), but there was no significant difference in the CRP level on discharge (*p*=0·74). Compared with single-RP-DC patients, multiple-RP-DC patients had longer hospitalization periods (median: 24 *vs* 18 days, *p*=0·02) and viral RNA duration times (median time from onset to last RP: 65 *vs* 33 days, *p*<0·0001), but had no significant differences in their other demographic or clinical characteristics.

During readmission, 67 of 93 RP-DC patients (72%) had no symptoms, while 26 (28%) had mild symptoms, including slight cough (18/93 [19%]) and chest tightness (3/93 [3%]). One patient (male, 12 years old) had a brief fever (temperature: 37.5 °C) for one day. Routine blood tests showed elevated interleukin 6 levels in one patient (male, 62 years old); all other patients had normal levels. Chest CT revealed that 18 (19%) patients had no pneumonia lesions and the lung lesions of the remaining 75 patients were improved (68/93, 73%) or unchanged (7/93, 8%) from first discharge. There were no significant clinical symptom differences between single- and multiple-RP-DC patients during readmission.

Seventy-one (76%) RP-DC patients were identified by only positive nasopharyngeal swab results, 14 (15%) by only positive anal swab results, and 8 (9%) by positive results for both specimen types. All tested blood specimens (147/147) from RP-DC patients were SARS-CoV-2 RNA negative. The median Ct values of N and Orf1ab genes were 35 (95% CI: 35–36) and 36 (95% CI: 36–37), respectively, which are significantly higher than the corresponding values at disease onset (N gene median Ct: 31, 95% CI: 29–31; Orf1ab gene median Ct: 31, 95% CI: 30–32, *p*<0·0001; Figure 2c). Furthermore, RP-DC patient viral RNA levels ranged from 1·9 to 5·7 log_10_ copies/mL (median: 3·1, 95% CI: 3·0–3·2), which was significantly lower than the corresponding values at disease onset (median: 4·5 log_10_ copies/mL, 95% CI: 4·3–4·8, *p*<0·0001; Figure 2d), indicating low viral RNA levels in RP-DC patients. Most (89/93; 96%) RP-DC patients had a maximum viral RNA level of <5 log_10_ copies/mL. There was no significant difference in viral RNA levels between patients of different demographic and clinical categories, between single- and multiple-RP-DC patients, or between positive nasopharyngeal and anal swab specimens (*p*>0·05, appendix Figure S3). There was a significant negative correlation between discharge time and viral RNA level (*R*=0·20, *p*=0·002; Figure 2a), and the viral RNA level of multiple-RP-DC patients showed a declining trend as the number of RP detections increased (Figure 2e).

To investigate the antibody responses of RP-DC and NRP-DC patients, their SARS-CoV-2-specific anti-RBD IgM, IgG, IgA, total antibody, and NAb were assessed. A total of 499 serum specimens were obtained from 78 RP-DC patients (289 specimens, 1–9 specimens/patient) and 94 NRP-DC patients (210 specimens, 1–6 specimens/patient) within 14 weeks post-discharge (within 17 weeks post-disease onset). The IgM, IgG, IgA, total antibody, and NAb seropositivity rates at first post-discharge sampling (median: 24 days post-discharge) in RP-DC patients were 37% (29/78), 99% (77/78), 62% (48/78), 99% (77/78), and 88% (69/78), respectively, with a median NAb titre of 1:32 (95% CI: 1:16–1:32), which were not significantly different (*p*>0·05) from those of NRP-DC patients (50% [47/94], 98% [92/94], 50% [47/94], 99% [93/94], and 92% [77/84], respectively; median NAb titre: 1:16, 95% CI: 1:16–1:32). For RP-DC patients whose specimens were collected on the day of RP detection, these rates were 38% (18/48), 98% (47/48), 63% (30/48), 100% (48/48), and 91% (39/43), respectively, with a median NAb titre of 1:32 (95% CI: 1:16–1:32).

We further quantitatively investigated the RP-DC and NRP-DC patient antibody levels during different sampling periods. Seventy five percent of RP-DC patients were identified during their two-week quarantine; no significant differences from NRP-DC patients were identified in specimens from this period (Figure 3a). During our entire sampling period (3–17 weeks post-disease onset), no significant weekly differences were identified, except the IgM and total antibody level in week 3 and IgM level in weeks 6–8 (*p*<0.05, Figure 3b). Specifically, one (1%) and five (6%) RP-DC patients were negative for IgG and NAb, respectively, which is not significantly different (*p*>0·05) from NRP-DC patients (IgG-negative: 3% [3/94]j; Nab-negative: 8% [7/84]). Furthermore, we compared the RP-DC patient antibody levels on the day of RP detection and within one week before and after RP detection (when patients were viral RNA negative); no significant differences were identified (Figure 3c). Together, these results indicate that the SARS-CoV-2-specific anti-RBD antibody levels (excluding IgM) are similar in RP-DC and NRP-DC patients and in RP-DC patients regardless of current RP detection. Additionally, there was a significant correlation between NAb titres and antibody levels (*R*>0·40, *p*<0·0001), particularly for IgG (*R*=0·73, *p*<0·0001) and total antibody (*R*=0·77, *p*<0·0001), which indicates that they may be alternative indicators of NAb titre (appendix Figure S4).

**Figure 3.**
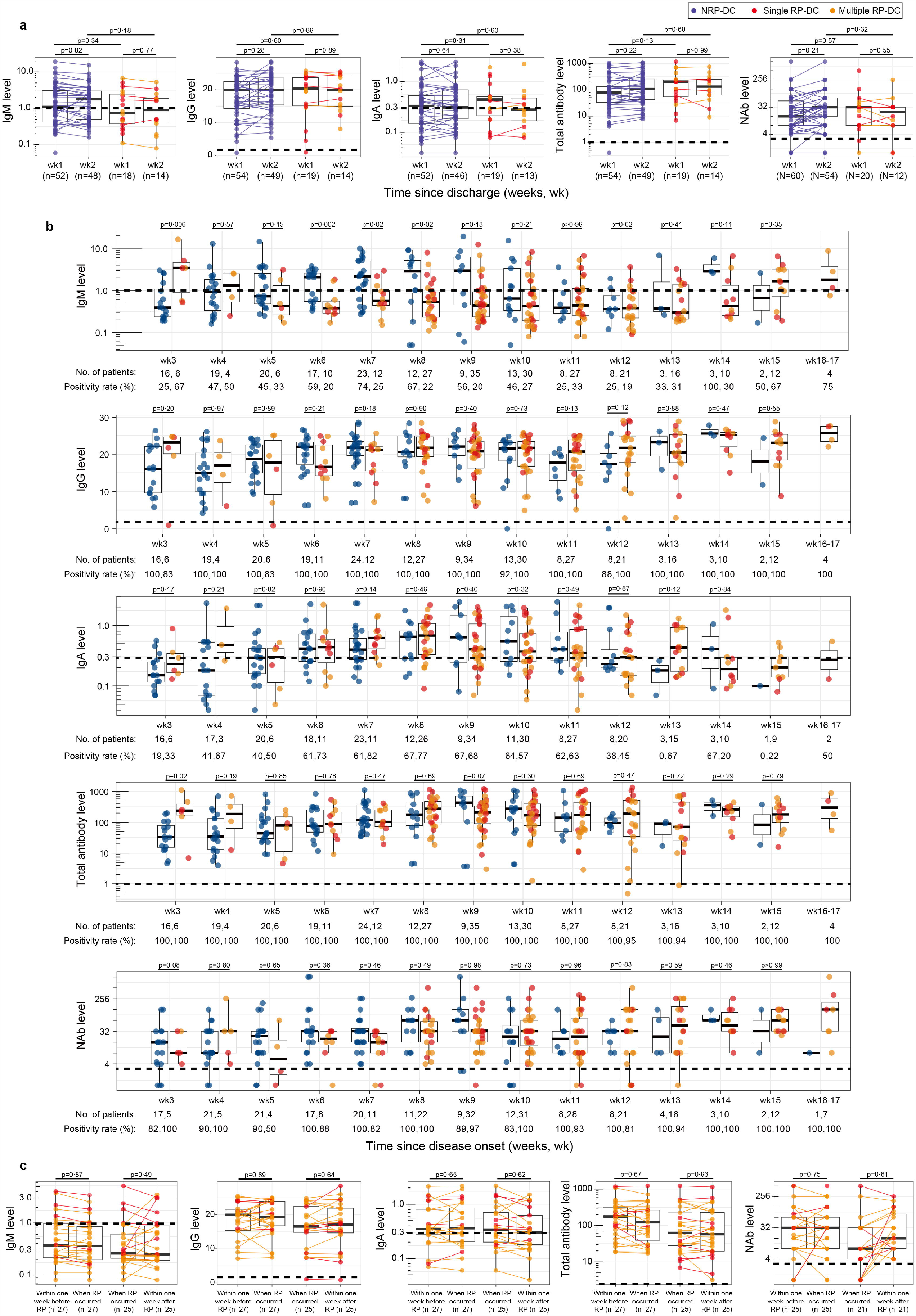
Serum SARS-CoV-2-specific antibody levels in RP-DC and NRP-DC patients. (a–b) Levels of antibody against SARS-CoV-2 surface spike protein receptor-binding domain in RP-DC and NRP-DC patients within two weeks post-discharge (a) or since disease onset (b). (c) Anti-SARS-CoV-2 surface spike protein receptor-binding domain antibody levels in RP-DC patients within one week before RP detection, at the time of RP detection, and within one week after RP detection. Blue, red, and orange points show NRP-DC, single-RP-DC, and multiple-RP-DC patients, respectively. Specimens from individual patients are linked by lines. Horizontal dotted lines indicate the positive detection threshold.

Virus isolation and WGS were performed to test whether live virus and/or complete viral genome, respectively, were detectable in RP-DC patients. Viral isolations of nine RP-DC patient nasopharyngeal specimens with representative Ct values (27–39, four specimens with a Ct value of <30 were included) were negative, as confirmed by testing the cell culture for SARS-CoV-2 RNA. WGS was successful for six of the nine specimens, but only genome fragments were obtained. The genome coverage of the specimens with the lowest Ct value (Ct: 27) was 55%, whereas the coverage of other specimens was <10%.

To assess whether RP-DC patients could spread the virus to close contacts, we conducted prompt epidemiological investigations of 23 RP-DC patients (identified during follow-up) on the day of RP detection, which identified 96 close contacts. None showed clinical symptoms during the two-week follow-up, and all had negative SARS-CoV-2 RNA test results; 20 were tested for serum SARS-CoV-2-specific anti-RBD antibodies (IgM, IgG, and total antibody), and the results were also negative. Notably, one paediatric RP-DC patient was identified at 90 days post-discharge, after being in school for 11 days, and all 1,200 of his candidate contacts (teachers and classmates) showed no clinical symptoms during fourteen-day observation and had negative results from SARS-CoV-2 RNA tests. As of July 10, no close or candidate contacts of RP-DC patients had become confirmed COVID-19 patients. Additionally, a retrospective investigation of the contact history of 154 COVID-19 patients after February 1 found that none were epidemiologically related to our RP-DC patients. These results provide direct evidence that RP-DC patients have a low viral transmission risk.

All RP-DC patients were re-discharged after obtaining negative SARS-CoV-2 RNA detection results during quarantine. As of July 10, none of our RP-DC patients had any further RP results from SARS-CoV-2 RNA tests, i.e. all were fully recovered. Among the 479 fully recovered COVID-19 patients, NAb titres were tested in 162 (84 NRP-DC and 78 RP-DC patients), 93% (151/162) of whom were NAb-positive with a median titre of 1:32. Notably, five patients developed detectable NAb during quarantine or follow-up, including three RP-DC and two NRP-RC patients, whereas 11 fully recovered patients remained NAb negative during our sampling period. Based on the reverse cumulative distribution curve principle^13^, we analysed the NAb titre distribution at the end of quarantine for 162 fully recovered COVID-19 patients (Figure 4). RP-DC and NRP-DC patients had similar NAb titre distributions. Although some patients had a high NAb titre (28% with NAb titre of ≥1:64), 60% of fully recovered patients had NAb titres of ≥ 1:32. Thus, this value could be used as a reference indicator for evaluating humoral responses to COVID-19 vaccine candidates in future clinical trials.

**Figure 4.**
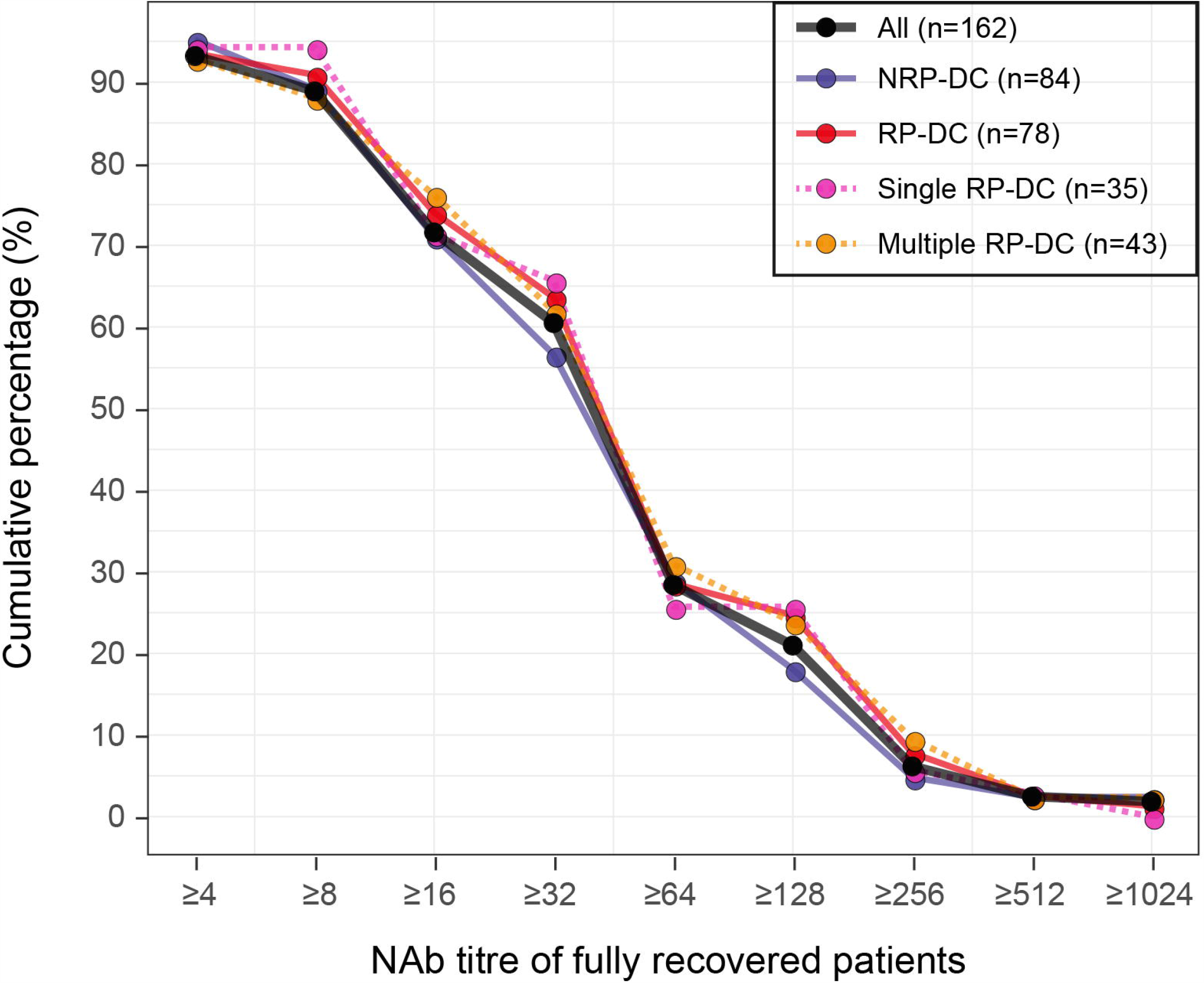
Reverse cumulative distribution curves of NAb titres in fully recovered patients. Colours show different types of patients.

## Discussion

To our knowledge, this is the first population-based study to comprehensively describe the viral RNA level and antibody response characteristics of RP-DC patients and evaluate their viral transmission risk. RP-DC patients were characterised by younger age, mild or absent symptoms, and no disease progression. They generally had low viral RNA levels but long viral RNA durations (up to 113 days post-disease onset). Although the prolonged presence of SARS-CoV-2 RNA in COVID-19 patients has been reported,^14, 15^ our results suggest that low levels of SARS-CoV-2 RNA persisted in some patients after both clinical recovery and initial viral-negative conversion. Except for IgM, no significant differences in antibody or NAb levels were identified between RP-DC and NRP-DC patients or in RP-DC patients over time (before, during, or after RP detection), suggesting that RP occurrence may not be related to humoral immunity. The low viral RNA levels and effective, long-lasting antibody responses in RP-DC patients, combined with the failed virus isolation, fragmented genome detection, and lack of close contact infections from these individuals, suggest that RP-DC patients pose a low risk of viral transmission. Furthermore, 60% of the fully recovered COVID-19 patients had NAb titres of ≥1:32; this value could be used to evaluate the humoral response in COVID-19 vaccine clinical trials.

By systematically monitoring SARS-CoV-2 RNA in DC patients during quarantine and follow-up, we found that RP-DC patients accounted for 19% of DC patients, which is close to most previous reports (15%–21%)^7, 9, 10^ but much higher than one recent report where 3% (23/651) of RP-DC patients were identified in a routine health check of DC patients^.16^ Considering that multiple negative RNA tests were also identified in our RP-DC patients, differences in detected RP-RC patient proportions may be related to the viral RNA testing frequency. However, in the context of systematic follow-up and testing, RP occurrence in DC patients is unlikely to be rare.

Although RP-DC patients have been observed by multiple independent researchers^6-10^ and government authorities, including the Korean CDC^,17^ the cause of RP occurrence remains unclear, and several hypotheses have been proposed. 1) RP might be due to false-negative SARS-CoV-2 RNA test results at discharge^.9, 18^ Here, in the 59% of RP-DC patients who had additional negative test results before their first RP result, the sampling and testing were performed by the same technician using the same kits, minimizing the likelihood of false-negative results. 2) RP could be due to post-discharge reinfection. Here, 75% of RP-DC patients were identified during quarantine, and those identified during follow-up did not report any contact with COVID-19 patients, making reinfection unlikely. 3) In people with low antibody levels or immunity, uneradicated virus could cause secondary infections.^19^ We did not detect significant differences in antibody levels between RP-DC and NRP-DC patients or in RP-DC patients over time, suggesting that humoral immunity may not be related to RP occurrence. Additionally, none of the RP-DC patients had immunodeficiency diseases, and there was no significant difference in steroid treatment between RP-DC and NRP-DC patients. However, more data are needed to verify the relationship between RP occurrence and immunity, especially regarding cellular immunity. 4) RP occurrence may be due to the shedding of ‘dead’ virus particles. This possibility is consistent with our negative virus isolation results. However, failed viral isolation does not confirm a lack of live virus; Wölfel and colleagues^20^ found that live virus cannot be successfully isolated when the viral load is below 10^6^ copies/mL. More sensitive live virus detection methods, such as identification of subgenomic messenger RNA^20^, are needed to prove this hypothesis. Based on our data from SARS-CoV-2 RNA testing on 2,589 clinical samples collected from February 18 to May 5, eleven RP-RC patients were identified ≥30 days post-discharge (maximum: 90 days post-discharge), and all patients had recovered; therefore, we propose that RP occurrence in DC patients is due to their intermittent and non-stable excretion of low levels of viral RNA. However, further studies on the mechanism of RP occurrence are needed.

Because SARS-CoV-2 RNA positivity does not necessarily translate to infectivity, we integrated multiple approaches to systematically evaluate the viral transmission risk posed by RP-DC patients. The viral RNA level can be a useful indicator for accessing transmission risk. Wölfel and colleagues^20^ proposed that patients with a viral load of <5 log_10_ copies/mL posed a low transmission risk based on virus isolation results. Here, 96% of RP-DC patients had a maximum viral RNA level of <5 log_10_ copies/mL (range: 1·9–5·7 log_10_ copies/mL). Four RP-DC patients had a maximum viral RNA level of >5 log_10_ copies/mL, linked with a possible risk of viral transmission. To assess whether RP-DC patients shed live virus, we attempted virus isolation on the four specimens with a viral RNA level of >10^5^ copies/mL and five representative specimens with lower viral RNA levels. All nine specimens produced negative results. The low viral RNA levels and negative virus isolation in samples from the RP-DC patients indicate that their transmission risk is low.

WGS can be used to identify viruses with specific mutations, the presence of which may identify reinfection from another source. However, we obtained only genome fragments from the RP-DC patient specimens after SARS-CoV-2-specific amplification, including the specimen with the lowest Ct value (Ct: 27, viral RNA level: 5·7 log_10_ copies/mL), which limited our further investigation. In comparison, Liu and colleagues^21^ found that sequencing reads can cover ≥90% of reference genomes with a Ct value of <30, irrespective of the amplification and sequencing approach. Although technique differences exist, the low genome coverage of RP-DC patient specimens suggests a low viral RNA level, further supporting the idea that RP-DC patients pose a low transmission risk.

The most effective way to assess the transmission risk of RP-DC patients is to conduct epidemiological investigations of their close contacts. When conducting epidemiological investigations on 790 close contacts of 285 RP-DC patients, the Korean CDC did not identify any infections^.17^ However, the possibility of asymptomatic infections in those contacts was not excluded through SARS-CoV-2 RNA testing and antibody testing. Here, not only did all 96 close contacts and 1,200 candidate contacts show no clinical symptoms, they also had negative SARS-CoV-2 RNA test results, and 20 of them had negative antibody results, suggesting there were no asymptomatic infections. As of June 10, no COVID-19 cases have been reported among those contacts. These findings directly support our conclusion that RP-DC patients pose a low transmission risk. Furthermore, the RP-DC patients had high and long-lasting NAb levels, suggesting that they can effectively clear virus, which further reduces their viral transmission risk.

Whether COVID-19 convalescent patients are protected against future SARS-COV-2 infections is largely unknown.^22, 23^ NAb play important roles in virus clearance and are considered vital for protection against viral disease. Among the 162 fully recovered RP-DC or NRP-DC patients who were tested for NAb, 93% (151/162) were NAb positive, with a median titre of 1:32, and their detectable NAb was maintained for up to 17 weeks post-disease onset, suggesting that most recovered patients obtained effective and long-lasting protection against future SARS-CoV-2 infection. Effective vaccines against SARS-CoV-2 infection are urgently needed to reduce the burden of COVID-19, and more than 120 candidate vaccines are currently being developing worldwide.^5, 24, 25^ NAb titres in recovered COVID-19 patients make ideal reference values to use as vaccine humoral immunogenicity endpoints in vaccine efficacy evaluations. Based on our finding that 60% of fully recovered patients had NAb titres of ≥1:32, future COVID-19 vaccine clinical trials might consider using this titre as a reference indicator for evaluating humoral responses.

Our study has several limitations. First, this was a single-centre study conducted on all DC patients from Shenzhen. Because there are differences in the discharge criteria and SARS-CoV-2 RNA testing methods among different cities and counties, our RP incidence needs to be verified by multicentre studies. Second, we collected only nasopharyngeal swab, anal swab, and serum specimens based on current sampling policies; other specimen types with generally higher viral loads, such as lower respiratory tract and sputum specimens, were not collected. Thus, the RP incidence in this study represents a conservative estimation. Third, the systemic collection of serum specimens started mid-study, and serum specimens from RP-DC patients during their hospitalization were not available, which limited further investigations on the antibody level dynamics of RP-DC patients. Finally, due to the strict management of DC patients, most DC patients were identified during quarantine and consequently had few close contacts. This study included the close contacts of only 23 RP-DC patients; larger scale epidemiologic studies are needed to further confirm the transmission risk posed by RP-DC patients.

In conclusion, our study found that intermittent detection of low levels of SARS-CoV-2 RNA in DC patients is not rare and that the timing of RP detection varies (up to 90 days post-discharge). The transmission risk posed by RP-DC patients is likely low. To better balance COVID-19 prevention and control with economic activities and to more effectively manage DC patients while minimizing the psychological impact on these individuals, we suggest that public health authorities should take a relatively relaxed approach to managing DC patients. However, the follow-up and personal protection of DC patients should be strengthened. Last, given that 60% of fully recovered patients had NAb titres of ≥1:32, this value may serve as a useful reference indicator for evaluating humoral responses to COVID-19 vaccine candidates in future clinical trials.

## Data Availability

All the data are available

## Contributors

RY, YL, SM, BL, ZR, and QH conceived and supervised this study. CY, MJ, XW, SF, HL, LZ, YJ, YZ, QC, CZ, LW, SW, WW, YL, HZ, and HYZ performed laboratory tests. XT, HL, JY, XL, ZZ, XR, XZ, TF, JX, YG, MW, and LL collected and organised data. CY, MJ, XW, QH, YC, and RY performed data and results interpretation. CY, XW, and MW conducted statistical analyses. CY, MJ, RY, and QH drafted the manuscript and figures. All authors reviewed and approved the final version of manuscript.

## Acknowledgments

This research was supported by Sanming Project of Medicine in Shenzhen (No. SZSM201811071), the China National Science and Technology Major Projects Foundation (No. 2017ZX10303406), Special Foundation of Science and Technology Innovation Strategy of Guangdong Province of China (No. 2020B1111340077), and a Shenzhen Committee of Scientific and Technical Innovation grant (No. JCYJ20180508152244835). We thank Prof. Fengcai Zhu from Jiangsu Center for Disease Control and Prevention, China, for useful advice on neutralisation antibody titre analysis and critical editing of the manuscript. We thank Prof. Ming Zeng from Shenzhen Kangtai Biological Products Co., Ltd., for critical editing of the manuscript. We thank Gillian Campbell, PhD, and Katie Oakley, PhD, from Liwen Bianji, Edanz Group China (www.liwenbianji.cn/ac) for editing the English text of drafts of this manuscript. We thank all the patients who consented to donate their data for analysis and the medical staff members who are on the front line of caring for patients.

